# Study Design Indexing in Transition: A Focused Comparison of manual NLM Indexing vs. Transformer-based Automated Models

**DOI:** 10.64898/2026.06.03.26354854

**Authors:** Puranjani Das, Jodi Schneider, Evan Mayo-Wilson, Halil Kilicoglu, Joe D. Menke, Dongin Nam, Kiran Ninan, Jean-Pierre Oberste, Ang Michael Troy, Xiangji Ying, Arthur W. Holt, Neil R. Smalheiser

## Abstract

**Objectives:** We sought to evaluate the accuracy and suitability of a transformer-based model (TM) for indexing clinical study designs, in comparison to National Library of Medicine (NLM) indexing.

**Materials and Methods:** For 2016 (manual NLM) and 2025 (automated NLM) published articles, we conducted a limited evaluation of the TM model, with articles that received the most confident predictions, with which NLM assignments disagreed. Ground truth was established through independent dual annotation for four prominent study designs: cohort, case-control, cross-sectional, and case report.

**Results:** For three designs (case-control, case report, cross-sectional), the articles having the top 100 predictive TM scores (when NLM failed to assign that design) were judged to exhibit that design in the great majority (86-100%) of cases. Conversely, the articles having the lowest 100 predictive TM scores (when NLM did assign the study design) exhibited the design only in relatively few (0-21%) of cases. The most confident predictions of the TM model were highly accurate and not redundant with automated NLM indexing; the exception was cohort studies articles, in which both TM and NLM labels showed high error rates of both omission and commission.

**Discussion:** TM may have value for identifying articles exhibiting study designs, which is important for clinical decision-making and evidence syntheses.

**Conclusion:** NLM’s cohort studies indexing is unreliable as a gold standard for building automated systems, warranting efforts to create a new manually annotated corpus.

**LAY SUMMARY:** Collecting all relevant evidence on a given clinical question can be important. However, the existing indexing in PubMed/MEDLINE is neither accurate nor comprehensive enough for many purposes. Our group has created an automated machine learning model for finding and indexing articles using important clinical study designs. We compared our indexing against the existing indexing to see whether our approach adds value. When our model is highly confident, and disagrees with PubMed/MEDLINE indexing, our model is significantly more accurate. However, for one study design (cohort studies), both approaches frequently made errors such as missing studies or mislabeling them. For this study design, the current PubMed/MEDLINE indexing is not reliable enough to serve as a standard for training future automated indexing systems. This work advances automatic detection of important clinical study designs and complements existing PubMed/MEDLINE indexing.

## BACKGROUND AND SIGNIFICANCE

MEDLINE is a major bibliographic database of biomedical and life sciences literature maintained by the U.S. National Library of Medicine (NLM)^1^, and is a core component of the PubMed search engine. Until recently, biomedical articles were indexed manually by professional curators, employing a standardized hierarchical Medical Subject Heading (MeSH) terminology^2^. The majority of terms represent biomedical entities (e.g., lungs) or geographical names (e.g., Bosnia), and the goal of indexers was to assign about 8-20 of the most important topics discussed in a paper. In contrast, a subset of terms represent Publication Types (e.g., review, letter, or editorial) and the goal of indexers was to assign these terms to articles that exhibited (not merely discussed) the given article type. The situation is more complicated with regard to clinical study designs, because some of the designs (e.g., cohort studies, case-control studies, and cross-sectional design) are listed as topical terms whereas others (e.g., randomized controlled trial (RCT), case reports) are listed as Publication Types [1]. Therefore, an article may be indexed with one of these three clinical study design terms if it either discusses **or** exhibits that design.

Study design indexing is used for retrieval by individual users such as physicians, researchers and the public; however, it is particularly crucial for collecting all relevant evidence on a given clinical question, as carried out by groups preparing systematic reviews (SRs) and other evidence syntheses. Generally a systematic review will only consider articles exhibiting specific designs, e.g., only randomized controlled trials, or including specific observational controlled studies such as cohort studies and case-control studies. In practice, however, SR groups have found that NLM indexing is neither accurate nor comprehensive enough for their purposes. Indeed, previous studies have revealed that manual NLM indexing has a high rate of errors of both omission (failing to assign terms) and commission (mis-assignment of terms) (reviewed in Smalheiser et al., 2026). Therefore, the initial retrieval of articles for a SR tends to bypass NLM indexing entirely, resulting in a very large initial set (e.g., > 5,000 articles) that requires manual triage screening to reduce to a much smaller number (e.g., 10-50 articles) for full-text scrutiny.

In order to create a more suitable indexing system for publication types and study designs (collectively, PTs), our group has created a series of automated machine learning-based models that assign probabilistic predictive scores (0<p<1) that a given article exhibits a given study design [3]. We first employed Support Vector Machine (SVM)-based models, first for assigning RCTs [4], then extended to 50 PTs. More recently, we have developed a more accurate transformer-based model, TM, that currently indexes 73 PTs [5]. By assigning predictive scores instead of TRUE/FALSE assignments, we allow users to adjust their retrieval for high precision or high recall; for example, setting a very low threshold (p<0.01) permits nearly perfect recall of RCT articles while removing approximately 64% of the non-RCT articles from the initial retrieval set, thus resulting in significant savings of effort during SR triage [6].

At the same time, NLM has gradually adopted automated indexing technologies [7]. First, systems such as the Medical Text Indexer (MTI)^3^ used natural language processing and machine learning to analyze article titles and abstracts and suggest appropriate MeSH terms to human indexers. Currently, the neural network-based MTIX model is used to index MeSH terms in MEDLINE articles [8] and automated models are employed for Publication Types as well [9].

## OBJECTIVES

We seek to evaluate the accuracy and suitability of our TM model for indexing publication types and particularly, for indexing clinical study designs, in comparison to NLM indexing. However, this is challenging for at least three reasons: First, to date, all automated systems have been trained and evaluated on manual NLM indexing assignments [10–15], which is itself subject to errors [2,16,17]. Second, our probabilistic predictive scores take into account atypical articles that are difficult to assign, and can be converted to TRUE/FALSE assignments in different ways depending on the needs of users, while NLM indexing is categorical. Third, our goal (to tag articles only that exhibit a given design) differs from NLM indexing which tags articles that both discuss as well as exhibit that design.

In the present paper, we have accordingly carried out a limited evaluation of the TM model that focuses only on the articles that received the most confident predictions, that is, the highest scores that are almost certainly TRUE and the lowest scores that are almost certainly FALSE, but which disagreed with NLM assignments. The evaluation was limited to these cases because extreme disagreements between highly confident model predictions and NLM indexing could reveal systematic differences in indexing criteria or recurring model errors. The evaluation was performed both for articles published in 2016 (when NLM decisions were manual) and in 2025 (when all NLM decisions were automated). To establish ground truth, we employed dual annotators to index the articles independently, based on written definitions.

## MATERIALS AND METHODS

Figure 1 shows the 3 different phases of the study: selecting a subset of NLM-indexed articles to analyze, experts manually annotating study designs, and finally analyzing patterns of discrepancies compared to NLM indexing.

**Figure 1:**
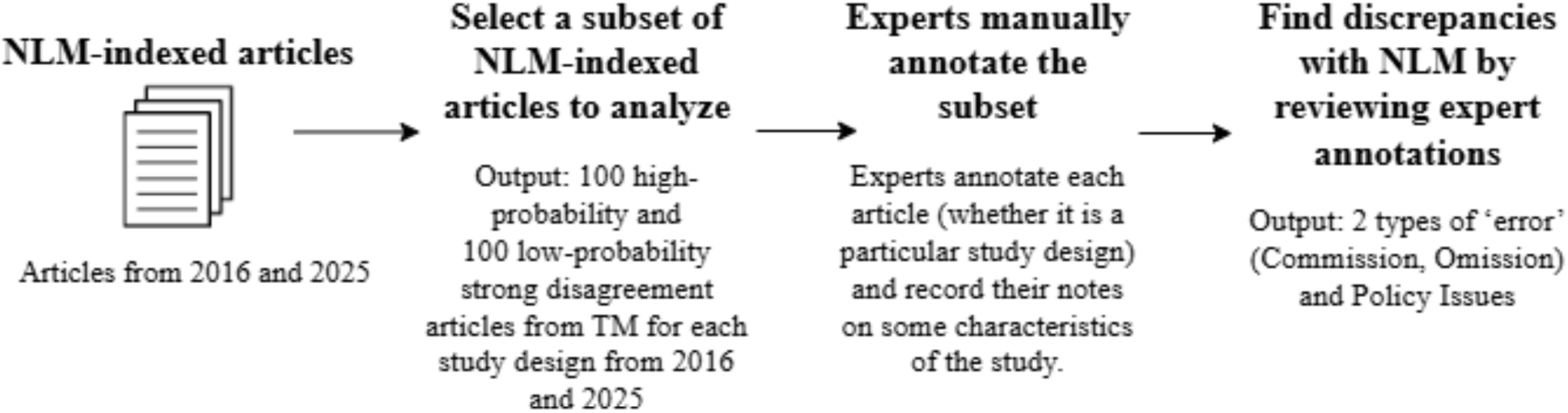
Overview of our study. [ALT TEXT: Figure 1: Flowchart showing an overview of the three-step process for comparing TM and NLM indexing of articles: selecting article subsets, experts manually annotating study designs, and identifying indexing errors and policy issues.]

Initially, we chose five prominent study designs to examine, including several terms listed as Publication Types (systematic review, case reports) and several observational study types listed as topical MeSH terms (cohort studies, case-control studies, and cross-sectional design) that are often included in evidence syntheses. In preliminary studies, we found that Systematic Review had excellent indexing agreement across all methods, and so only the latter four study designs were examined further in detail. Note that capitalized terms, e.g. Cohort Studies, are regarded as indexing terms, whereas when non-capitalized, refer to articles exhibiting the study design.

### TM indexing model

TM is a single multi-label model [5] that fine-tunes SPECTER2-base [18], a BERT-based encoder pre-trained on biomedical literature using citation-informed contrastive learning. TM uses title, abstract, and metadata (e.g., numbers of references), with asymmetric loss, contrastive learning, and label smoothing applied during fine-tuning to improve robustness and calibration [5]. The model operates independently of existing NLM-assigned MeSH terms and publication types (i.e., they are not included within the model’s input). TM gives calibrated probabilistic scores between 0 and 1, indicating the predicted probability that a given article is of a particular study design. Probabilistic scores for a given label can be converted to binary values using any chosen threshold, where scores above the threshold are classified as true and those below are false. There is no single threshold that is appropriate for all literature uses as users may prioritize specificity or sensitivity in retrieving literature given the different goals of different users, such as finding or excluding all items of a given study design. By default, TM selects thresholds per-label that maximize the F1 score for each study design using a held-out validation set [5].

### Dataset of 2016 and 2025 articles

For the NLM’s human indexing system, we collected the articles from 2016, the most recent year when articles were indexed ‘fully’ by human experts (albeit with suggestions provided by an early automated system) [7]. We employed records indexed by a human but excluded curated indexing, in which a human reviews (and possibly modifies) an algorithm’s results^4^, because it is not a true test of either human or automated NLM indexing. The following query was used to fetch the articles from 2016 [retrieved on 22 January 2026]: `2016[dp] AND english[la] AND hasabstract AND (medline[sb] NOT (indexingmethod_curated OR indexingmethod_automated))`. For the 2016 manual subset, we initially retrieved 708,912 articles. Because the TM model was trained on publications from 1987-2023, we excluded 58,278 articles from 2016 in the training set to prevent data leakage, as well as 251 articles that had fewer than 25 words in title and abstract combined. Thus, the final article list for the 2016 manual subset had 650,383 records.

For the NLM’s automated indexing system, we collected the articles from 2025 as NLM completed the transition to fully automated indexing in 2022, allowing new MEDLINE citations to be indexed without routine human intervention [7]. The following query was used to fetch the articles from 2025 which are automatically indexed [retrieved on 22 January 2026]: `2025[dp] AND english[la] AND hasabstract AND medline[sb] AND indexingmethod_automated.` For the 2025 automated subset, we initially retrieved 784,652 articles. Of these, we found that six showed a Redirection

Notice^5^ indicating that the article had been deleted as a duplicate of another article, and one article (PMID:40912927) could not be found at all. 2,357 articles had less than 25 words in title and abstract combined, hence we removed them. In addition, we excluded 14 articles which lacked MeSH terms altogether. After these removal steps, the final 2025 automated subset included 782,288 articles.

### Collecting articles with highly confident TM scores that disagree with NLM indexing

For the 2016 and 2025 articles collected as above, we computed their TM predictive scores for four key study designs: case-control studies, case reports, cross-sectional studies, and cohort studies. For each study design and for each year (2016, 2025), we examined the articles having the 100 highest predictive TM scores that were NOT assigned the same label by NLM. Conversely, we also examined the articles having the 100 lowest predictive TM scores, that WERE assigned by NLM.

As a default, we normally convert the predictive scores to binary TRUE/FALSE decisions using the thresholds giving the optimal F1 performance for each study design [5]. Note, however, that the very highest TM scores (almost all > 0.9) would all predict TRUE and the very lowest TM scores (almost all < 0.1) would all predict FALSE (Table 1), regardless of the decision threshold values chosen over an extremely wide range. NLM indexing decisions were scored as TRUE if NLM assigned a given article the given study design, and FALSE if they did not.

**Table 1:** Mean score and range of the top 100 and lowest 100 TM model predictive scores.

| Study Design | Year | Mean Score | Score Range |
| --- | --- | --- | --- |
| **TOP 100** |  |  |  |
| Cohort studies | 2016 | 0.883 | 0.861 - 0.952 |
| Cohort studies | 2025 | 0.843 | 0.82 - 0.907 |
| Case-control studies | 2016 | 0.943 | 0.925 - 0.974 |
| Case-control studies | 2025 | 0.922 | 0.888 - 0.98 |
| Case reports | 2016 | 0.978 | 0.97 - 0.99 |
| Case reports | 2025 | 0.972 | 0.959 - 0.991 |
| Cross-sectional studies | 2016 | 0.987 | 0.984 - 0.994 |
| Cross-sectional studies | 2025 | 0.988 | 0.986 - 0.994 |
| **LOWEST 100** |  |  |  |
| Cohort studies | 2016 | 0.002 | 0.0 - 0.004 |
| Cohort studies | 2025 | 0.035 | 0.003 - 0.070 |
| Case-control studies | 2016 | 0.001 | 0.0 - 0.003 |
| Case-control studies | 2025 | 0.091 | 0.0 - 0.151 |
| Case reports | 2016 | 0.006 | 0.0 - 0.013 |
| Case reports | 2025 | 0.011 | 0.0 - 0.043 |
| Cross-sectional studies | 2016 | 0.003 | 0.0 - 0.006 |
| Cross-sectional studies | 2025 | 0.062 | 0.002 - 0.133 |

Table 1 shows the range of TM predictive scores for these articles. In most cases, the top 100 scores were all above 0.9, and the lowest 100 scores were below 0.1, indicating very high-confidence TRUE or FALSE predictions. Some of the scores in the Cohort Studies and Case-Control Studies subgroups fell slightly outside this range; this agrees with previous evaluations (and data presented here) that have indicated that these study designs are more difficult to index accurately than the others [3,5].

### Manual annotation and analysis

For each of the articles selected, two annotators were given a spreadsheet with the title and abstract and independently determined whether the study design indexing term should be applied, based on a written definition for each study design (see Supplementary File). If the title and abstract did not suffice to make a clear decision, annotators were allowed to access the full-text (using a link provided with the annotation materials). They were instructed not to be influenced by other cues such as article types assigned by NLM or showing on the journal’s website. Cohort studies and case-control studies were annotated by an associate professor of epidemiology (EMW) an assistant professor of epidemiology (XY), and two PhD students in epidemiology (JPO and KN); case reports and cross-sectional studies were annotated by a professor in psychiatry (NRS) and a bachelor’s student double majoring in Art History and Business Management (AMT) with prior biomedical annotation experience. Decisions were reconciled by discussion, but we kept track of the extent and type of initial disagreements. In the analysis phase, authors PD and JS looked at annotators’ comments on individual articles to investigate possible policy disagreements, such as definitional differences.

## RESULTS

### Independent manual evaluation of disagreements

For 3 of the 4 study designs (case reports, case-control studies, and cross-sectional studies), the vast majority (86-100%) of articles in the top 100 were judged by the independent annotators to exhibit that design, despite not having been indexed as such by NLM (Table 2). This was true both for 2016 and 2025 articles. The disagreements do not appear to be explained by indexing policy differences (Table 2), but instead, these appear to be NLM errors of omission. In this subset, TM only made 0-14% errors of commission (Table 2).

**Table 2.** Manual evaluation of the articles having the top 100 TM model scores for each study design and both years. Also indicated is whether we interpret differences as a policy difference, a TM error, or an NLM error.

| **Annotators' Manual Assessment** |  | **Article is SR, Meta-Analysis or Review** | **Annotators disagree with TM prediction** | **Annotators uncertain** | **Annotators agree with TM prediction** | **Total** |
| --- | --- | --- | --- | --- | --- | --- |
| **Interpretation** |  | **policy difference** | **TM error** |  | **NLM error** |  |
| **2016** | Cohort study | 1 | 32 | 0 | 67 | 100 |
|  | Case-control study | 1 | 13 | 0 | 86 | 100 |
|  | Case report | 1 | 1 | 3 | 95 | 100 |
|  | Cross-sectional study | 0 | 1 | 0 | 99 | 100 |
| **2025** | Cohort study | 0 | 41 | 1 | 58 | 100 |
|  | Case-control study | 0 | 5 | 0 | 95 | 100 |
|  | Case report | 0 | 0 | 0 | 100 | 100 |
|  | Cross-sectional study | 0 | 0 | 0 | 100 | 100 |

The situation is more complex with regard to cohort studies articles: NLM showed a high rate of errors of omission (67% in 2016 and 58% in 2025) but TM also showed a high rate of mis-assignments (32% in 2016 and 41% in 2025) (Table 2).

Conversely, as shown in Table 3, for all study designs, almost all (96-100%) of 2016 articles in the lowest 100 subset were judged NOT to exhibit that design. Thus, the TM model FALSE predictions were reliably accurate (error rate of 0-4%) in contrast to NLM manual indexing, which showed an error rate of 20-48% for that subset (Table 3). For articles indexed in 2025 (via the automated NLM MTIX algorithm), case reports and case-control studies also showed a low TM error rate (2-15%) and high NLM error rate (48-74%).

**Table 3.**
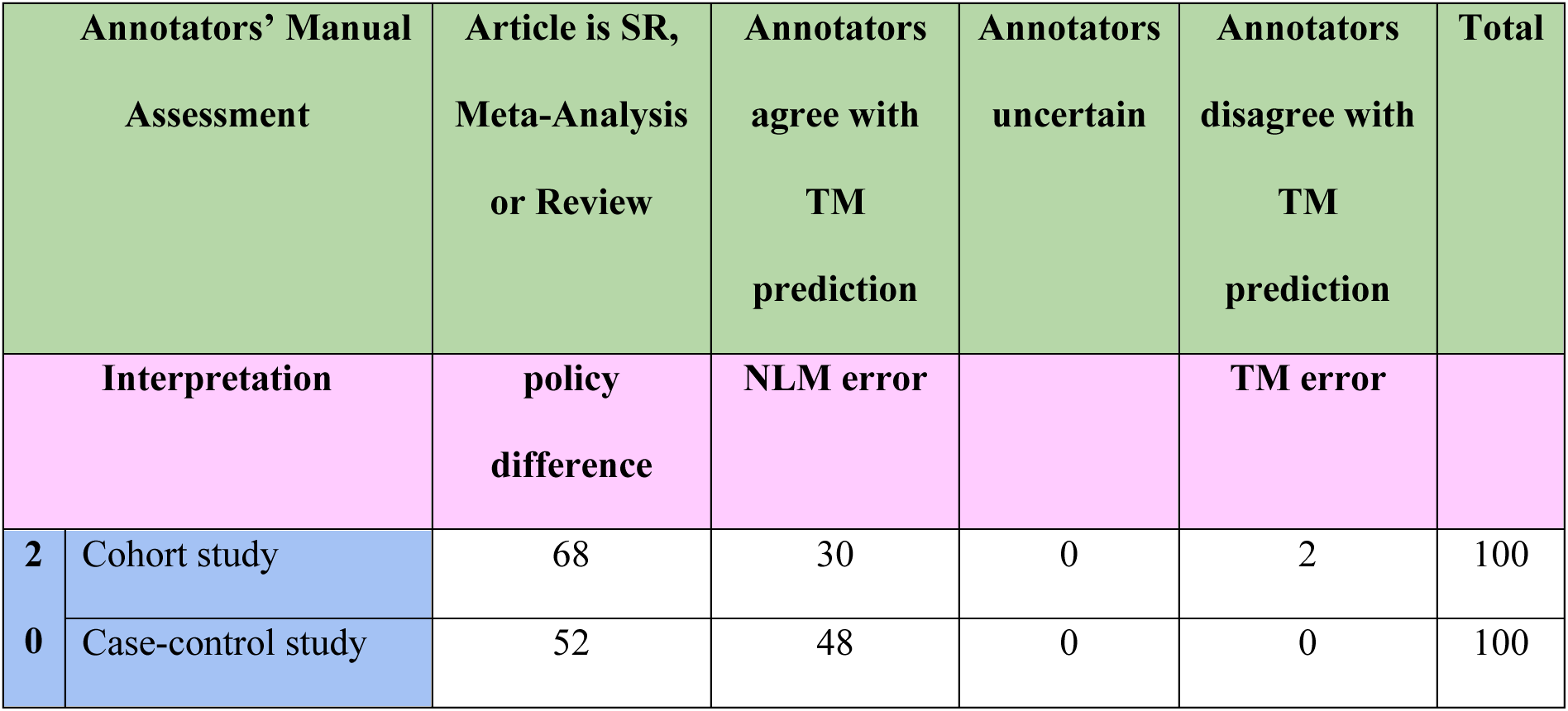
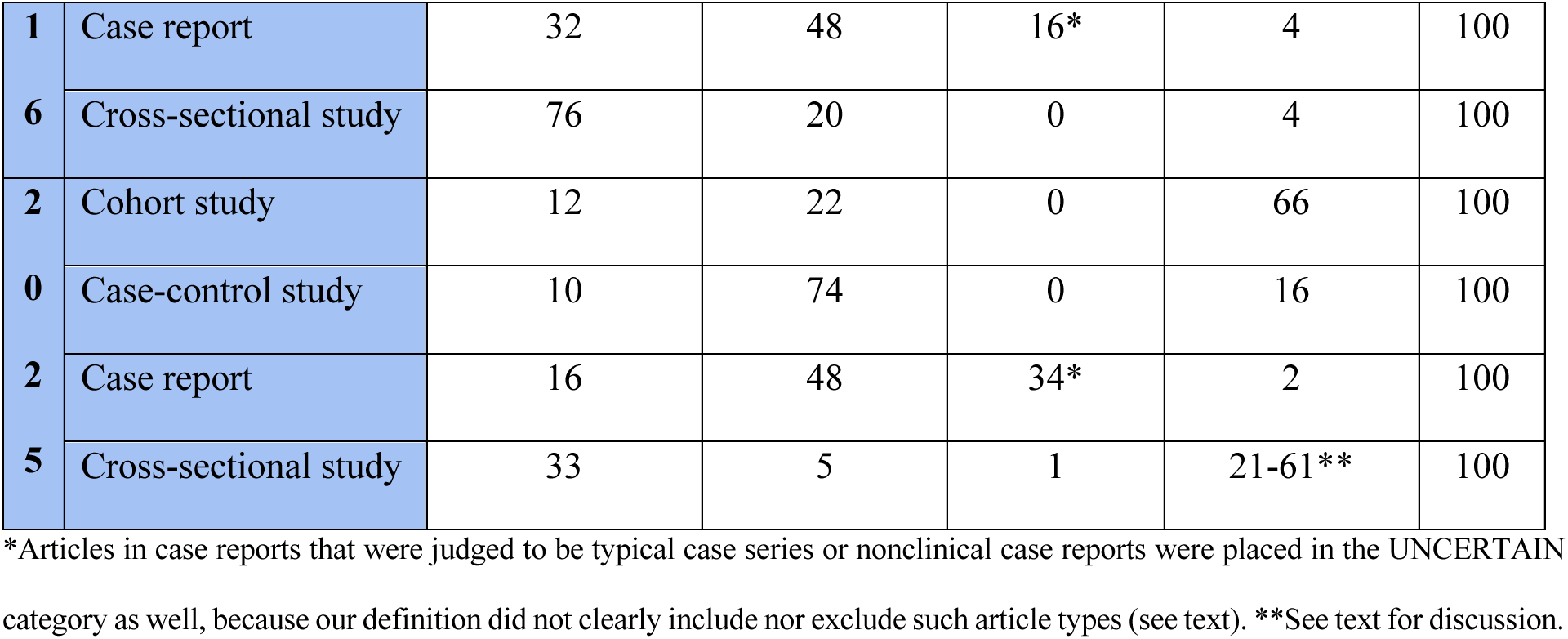
Manual evaluation of the articles having the lowest 100 TM model scores for each study design and both years. Also indicated is whether we interpret differences as a policy difference, a TM error, or an NLM error.

Surprisingly, of 2025 articles having very low TM predictive scores for cross-sectional studies but indexed as such by NLM, annotators marked 61% as correctly having that design (Table 3, double asterisks). Upon further analysis, we found that annotators noted that 40 of these 61 articles exhibited longitudinal designs. In fact, TM predicted that 51 of the total 100 articles in this subset satisfied the Longitudinal Studies design, and NLM assigned dual Cross-sectional Studies AND Longitudinal Studies terms to 52 of the 100 articles. Generally, longitudinal designs and cross-sectional designs are contrasting and typically quite different, and across all PubMed articles, only 9.3% of longitudinal studies are dually-indexed as cross-sectional. Apparently TM tends to assign Longitudinal Studies to such articles that also include some cross-sectional data, which appears to be an uncommon or edge case. If one regards that as a policy issue rather than simple mis-assignment error, then the TM model error rate should be more accurately estimated as (61-40)/100 = 21%.

Cohort studies was again a complex situation in 2025, wherein TM showed a high error rate (66%) and NLM also showed a significant error rate as well (22%, Table 3).

### Analysis of NLM and TM indexing errors

Beyond simply tabulating error rates as presented above, we also examined annotator notes to shed light on why TM and NLM made different assignments.

In the top 100 TM predictive score subset (Table 2), the TM model mis-assigned Cohort Studies more often than any of the other designs studied. Most of the TM errors (26 of 32 articles in 2016 and 18 out of 41 articles in 2025) were judged by the annotators to be cross-sectional studies instead. Although we can only speculate why the cross-sectional studies were mis-assigned, the word “cohort” is sometimes used to describe a group of people rather than a study design. Of the misclassified studies, 59% and 41% included the term “cohort” in the title and abstract, respectively. Interestingly, the automated NLM indexing also made similar mis-assignment errors: 13 of the 22 NLM errors in 2025 were judged by the annotators to be cross-sectional studies (Table 3).

In the lower 100 TM predictive score subset (Table 3), for all four of the study designs, NLM assigned the study design term to many systematic reviews, meta-analyses and other reviews that included or discussed articles having that study design (Table 3). This is consistent with their guidance to assign articles that **discuss** the study design, as well as those exhibiting the design to publication types not in MeSH like case-control studies or cohort studies. We did not count these as errors; instead, we regard these as policy differences between NLM and our own definitions (see Supplementary File) which only assign a term to articles that **exhibit** the given design.

Less clear is how to regard the articles that NLM marked as Case Reports that were not typical clinical case reports, but instead were case series articles or nonclinical case reports (Table 3, asterisks). On the one hand, case series do broadly satisfy the MeSH definition: “Clinical presentations that may be followed by evaluative studies that eventually lead to a diagnosis.” However, NLM is not very consistent in this regard, as about half of typical case series articles are **not** indexed as Case Reports by NLM [19]. As well, NLM assigning Case Reports to nonclinical case reports arguably is an error because a different MeSH term is applicable to most such studies (Organizational Case Series) [20]. We did not mark case series articles nor nonclinical case reports as errors, but placed them under UNCERTAIN as they were neither clearly included nor excluded by NLM nor by our written definition (see Supplementary File).

### 2016 manual NLM indexing vs. 2025 automated NLM indexing

Across the four study designs studied here, the transformer-based TM model shows consistently higher overall agreement with NLM in 2025 than 2016. This may not seem surprising, given that NLM reported using automated indexing in 2025 [8,21]. Yet our data indicate that, for articles that received high-confidence predictions by the TM model, the number of NLM errors did not change appreciably between 2016 and 2025.

In the top 100 subset, the NLM error rates were about the same in 2016 and 2025; if we sum up errors over all four designs, NLM error rate of omission in 2016 = (65+86+95+99)/400 = 86.25%, in 2025 = 87% (Table 2). In the lowest 100 subset (Table 3), the NLM error rate of commission was stable over time as well: in 2016 = 36.5% and in 2025 = 37.25%.

Similarly, in the top 100 subset, the TM error rate of commission was much less than NLM but still stable over time: in 2016 = 11.75%, and in 2025 = 10.5%. In the lowest 100 subset, the TM error rate of omission in 2016 = 2.5% and in 2025 = 26.25%. Overall, TM had much lower error rates than NLM which was true not only relative to manual NLM indexing, but to automated NLM indexing assignments as well.

## DISCUSSION

Attempting to compare the transformer-based TM indexing model against NLM indexing, over the entire set of PubMed articles, using independent manual annotators to establish ground truth, would be a daunting task. Therefore, we sought to focus attention on the most highly confident predictions of the TM model (both confident TRUE predictions and confident FALSE predictions), against both manual and automated NLM indexing decisions, and focused on four prominent study designs: one publication type (Case Reports) and three study designs (Cohort Studies, Case-Control Studies and Cross-Sectional Studies) that are frequently utilized by teams conducting evidence syntheses.

When the TM model gave its highest predictive scores to case reports, case-control studies, or cross-sectional studies, but NLM did not assign the study design, independent annotators judged that the TM model was correct in the great majority of cases (Table 2). This was true comparing the TM model both against manual NLM decisions (2016 articles) and against the automated NLM indexing (2025 articles). Similarly, when the TM model gave its lowest predictive scores to case reports, case-control studies, or cross-sectional studies, but NLM **did** assign the study design, annotators judged that the TM model was correct in the great majority of cases (Table 3).

Cohort studies articles were an exception insofar as both TM and NLM had relatively high error rates (Tables 3 and 4). On the one hand, NLM itself had a high error rate in assigning Cohort Studies, with 30% (2016) to 22% (2025) mis-assigned in our evaluation set. Apart from this, 68% (2016) to 12% (2025) of the articles marked by NLM as Cohort Studies consisted of systematic reviews and other reviews that discussed cohort studies. At the same time, the TM model – which was trained on the manual decisions of NLM curators but excluded review articles from training examples [5] – exhibited error rates for cohort studies higher than for the other study designs studied (Tables 3 and 4), and our previous global evaluation of the TM model vs. manual NLM indexing reported relatively low performance for cohort studies compared to most other publication types and study designs [5]^6^. As for **why** cohort studies should be so difficult to index accurately, both by TM and NLM, further research is needed, but a simple hypothesis is that the word “cohort” appears in many articles to describe a group of subjects in a diversity of contexts, requiring more than the usual effort to discern whether the study actually satisfies the definition of a cohort design. It is also likely that crucial methodological details are contained within the Methods section of full-text and often not clearly explained in the abstract.

These findings indicate that neither manual nor automated NLM indexing provides a reliable gold standard for indexing Cohort Studies, and warrant further efforts to develop independent manual corpora for both training and evaluation to improve the performance of automated indexing systems, especially for cohort studies but possibly for case-control studies as well, which exhibited high NLM error rates (Tables 3 and 4). These corpora needs to be large enough to be representative of the study design, and to deal not only with the most “typical” articles but a range of variations encountered in the literature.

In two previous publications, we carried out extreme disagreement analyses with experimental design similar to the present study. When the RCT Tagger tool was evaluated relative to manual NLM indexing, we found that NLM had relatively small error rates of omission (3%) and commission (7%) [4]; for that reason, we did not feel that it was warranted to re-examine RCTs here. The Multi-Tagger tool was evaluated relative to manual NLM indexing for three of the same study designs investigated here (case-control, cohort, and cross-sectional) [3] with results similar to those described in the present paper. However, we felt that it was important to repeat this evaluation for TM before publicly disseminating its predictive scores – and especially important to verify that highly confident TM predictions maintain their accuracy for the most recent biomedical articles, and that they hold their own against assignments indexed by the current automated MeSH system.

Our study has several limitations. We only examined the articles having the most confident TM scores which disagreed with NLM indexing, so the findings do not generalize to calculating the overall error rates of either TM or NLM over the entire literature. Also, study design definitions were either taken directly from the official MeSH definition, or consistent with it (see Supplementary File); however, we did not have access to the internal NLM curator guidelines. Although we believe that our definitions are non-controversial, they do not take into account all possible edge cases (e.g., how to index nonclinical case reports, or how to index longitudinal studies that include some cross-sectional data). Finally, we only examined four (albeit important) clinical study designs, so the results do not necessarily generalize to all publication types and study designs.

## CONCLUSION

In our dataset of four study designs, we find that highly confident TM model predictions are highly accurate in identifying true positives and true negatives, and – when they disagree with manual or automated NLM decisions – the TM model is significantly more accurate. The TM model advances automated PT tagging and complements NLM tagging systems.

## SUPPLEMENTAL MATERIAL

A supplementary file, containing definitions and annotator notes for manual examination of PubMed articles, is available with the journal. Separately, the protocol for this study was deposited at the Open Science Foundation (OSF) registry at https://osf.io/mbqsc/overview

## DATA AVAILABILITY STATEMENT

Data is available from the Illinois Data Bank at https://doi.org/10.13012/B2IDB-8307201_V1 [22] with:

- Spreadsheets containing manual annotations for each of 4 study designs before and after reconciliation: cohort studies, case-control studies, case reports, cross-sectional studies.
- Inter-annotator initial agreement for each study design and for each year, before reconciliation.

## CONTRIBUTORSHIP STATEMENT

- Ang Michael Troy: Data Curation
- Arthur W. Holt: Data curation, Formal analysis, Resources, Writing–review & editing
- Dongin Nam: Data curation, Resources, Writing–review & editing
- Evan-Mayo Wilson: Data Curation, Investigation, Writing–review & editing
- Halil Kilicoglu: Funding acquisition, Writing–review & editing
- Jean-Pierre Oberste: Data Curation
- Jodi Schneider: Conceptualization, Data curation, Formal analysis, Funding acquisition, Investigation, Methodology, Supervision, Writing–original draft, Writing–review & editing,
- Joe Menke: Writing–review & editing
- Kiran Ninan: Data Curation, Writing–review & editing
- Neil Smalheiser: Conceptualization, Data curation, Formal analysis, Funding acquisition, Investigation, Methodology, Supervision, Writing–original draft, Writing–review & editing
- Puranjani Das: Conceptualization, Data curation, Formal analysis, Investigation, Methodology, Writing–original draft, Writing– review & editing
- Xiangji Ying: Data Curation, Writing–review & editing

## Footnotes

1 https://www.nlm.nih.gov/medline/medline_home.html

2 https://www.nlm.nih.gov/mesh/meshhome.html

3 https://datadiscovery.nlm.nih.gov/Terminology/Medical-Text-Indexer-MTI-/ivwm-qssp/about_data

4 https://www.nlm.nih.gov/pubs/techbull/ja18/ja18_indexing_method_beta.html

5 For example, https://pubmed.ncbi.nlm.nih.gov/40774867/

6 See associated GitHub https://github.com/ScienceNLP-Lab/MultiTagger-v2/blob/main/AMIA/README.md

## REFERENCES

1. Module 2— MeSH Records, Descriptors, and Qualifiers (Overview). https://www.nlm.nih.gov/tsd/cataloging/trainingcourses/mesh/mod2_030.html (accessed 28 May 2026)

2 Smalheiser NR, Menke JD, Holt AW, et al. Goals and strategies for the indexing of publication types and study designs. Database (Oxford). 2026;2026:baag039. doi: 10.1093/database/baag039

3 Cohen AM, Schneider J, Fu Y, et al. Fifty ways to tag your PubTypes: Multi-tagger, a set of probabilistic publication type and study design taggers to support biomedical indexing and evidence-based medicine. medRxiv. 2021;medRxiv:2021.07.13.21260468. doi: 10.1101/2021.07.13.21260468

4 Cohen AM, Smalheiser NR, McDonagh MS, et al. Automated confidence ranked classification of randomized controlled trial articles: An aid to evidence-based medicine. J Am Med Inform Assoc. 2015;22:707–17. doi: 10.1093/jamia/ocu025

5 Menke JD, Kilicoglu H, Smalheiser NR. Publication type tagging using transformer models and multi-label classification. AMIA Annu Symp Proc. 2025;2024:818–27.

6 Schneider J, Hoang L, Kansara Y, et al. Evaluation of publication type tagging as a strategy to screen randomized controlled trial articles in preparing systematic reviews. JAMIA Open. 2022;5:ooac015. doi: 10.1093/jamiaopen/ooac015

7 U.S. National Library of Medicine. MEDLINE 2022 initiative: Transition to automated indexing. US NLM Tech Bull. 2021;443:e5.

8 U.S. National Library of Medicine. MTIX: The next-generation algorithm for automated indexing of MEDLINE. US NLM Tech Bull. 2024;457:e4.

9 Cid VH, Mork J. Enhancing automatic PT tagging for MEDLINE citations using transformer-based models. arXiv. 2025;arXiv:2506.03321v1. doi: 10.48550/arXiv.2506.03321

10 Du Y, Pan Y, Wang C, et al. Biomedical semantic indexing by deep neural network with multi-task learning. BMC Bioinformatics. 2018;19:502. doi: 10.1186/s12859-018-2534-2

11 Mao Y, Lu Z. MeSH Now: Automatic MeSH indexing at PubMed scale via learning to rank. J Biomed Semant. 2017;8:15. doi: 10.1186/s13326-017-0123-3

12 Peng S, You R, Wang H, et al. DeepMeSH: Deep semantic representation for improving large-scale MeSH indexing. Bioinformatics. 2016;32:i70–9. doi: 10.1093/bioinformatics/btw294

13 Rae AR, Mork JG, Demner-Fushman D. Convolutional neural network for automatic MeSH indexing. In: Cellier P, Driessens K, eds. Machine Learning and Knowledge Discovery in Databases. Cham: Springer International Publishing 2020:581–94.

14. Wang X, Mercer RE, Rudzicz F. MeSHup: Corpus for full text biomedical document indexing. In: Calzolari N, Béchet F, Blache P, et al., eds. Proceedings of the Thirteenth Language Resources and Evaluation Conference. Marseille, France: European Language Resources Association 2022:5473–83.

15 Wang X, Mercer R, Rudzicz F. KenMeSH: Knowledge-enhanced end-to-end biomedical text labelling. In: Muresan S, Nakov P, Villavicencio A, eds. Proceedings of the 60th Annual Meeting of the Association for Computational Linguistics (Volume 1: Long Papers). Dublin, Ireland: Association for Computational Linguistics 2022:2941–51.

16 Amar-Zifkin A, Ekmekjian T, Paquet V, et al. Algorithmic indexing in MEDLINE frequently overlooks important concepts and may compromise literature search results. J Med Libr Assoc. 2025;113:39–48. doi: 10.5195/jmla.2025.1936

17 Mork J, Aronson A, Demner-Fushman D. 12 years on – Is the NLM Medical Text Indexer still useful and relevant? J Biomed Semant. 2017;8:8. doi: 10.1186/s13326-017-0113-5

18 Singh A, D’Arcy M, Cohan A, et al. SciRepEval: A multi-format benchmark for scientific document representations. In: Bouamor H, Pino J, Bali K, eds. Proceedings of the 2023 Conference on Empirical Methods in Natural Language Processing. Singapore: Association for Computational Linguistics 2023:5548–66.

19 Shahidehpour A, Holt AW, Troy AM, et al. Creating an indexing scheme for case series articles. medRxiv. 2025;medRxiv:2025.12.19.25342712. doi: 10.64898/2025.12.19.25342712

20. Module 7—Publication Characteristics. https://www.nlm.nih.gov/tsd/cataloging/trainingcourses/mesh/mod7_060.html (accessed 30 April 2026)

21. U.S. National Library of Medicine. Frequently Asked Questions about Indexing for MEDLINE. https://www.nlm.nih.gov/bsd/indexfaq.html#terminology (accessed 10 March 2026)

22. Das P, Schneider J, Mayo-Wilson E, Nam D, Ninan K, Oberste J-P, Troy, AM, Ying X, Holt AW, Smalheiser NR. (2026): Comparison of NLM Publication Type Indexing in 2016 and 2025 vs. the Multi-Tagger Model vs. the Transformer Metadata Model [Dataset]. University of Illinois Urbana-Champaign Data Bank. doi:10.13012/B2IDB-8307201_V1

