## Supplementary File for "Study Design Indexing in Transition: A Focused Comparison of manual NLM Indexing vs. Transformer-based Automated Models"

### Definitions and annotator notes for manual examination of PubMed articles

#### 1. Cohort Studies

Definition taken directly from the official MeSH definition (https://meshb.nlm.nih.gov/record/ui?ui=D015331): Studies in which subsets of a defined population are identified. These groups may or may not be exposed to factors hypothesized to influence the probability of the occurrence of a particular disease or other outcome. Cohorts are defined populations which, as a whole, are followed in an attempt to determine distinguishing subgroup characteristics.

**Annotation Notes:**

1. If the study is classified as a cohort study, additionally assess whether:
   - The design is prospective
   - The unit of analysis is individual participants
   - The study evaluates the effect of an exposure observed after the start of the cohort on an outcome
2. For nested case-control studies and case-cohort studies, select "Yes" for both cohort study and case-control study

3. If the study is neither a cohort study nor a case-control study, select the appropriate option under other study design.
4. Please use the comment box sparingly. You may use it to:
5. Specify the study design if "Other" is selected, or
6. Note if the article appears to misclassify its study design.
7. Please use your own judgment rather than relying solely on the study's self-reported design.

### 2. Case-Control Studies

Definition taken directly from the official MeSH definition (https://meshb.nlm.nih.gov/record/ui?ui=D016022): Comparisons that start with the identification of persons with the disease or outcome of interest and a control (comparison, referent) group without the disease or outcome of interest. The relationship of an attribute is examined by comparing both groups with regard to the frequency or levels of outcome over time.

**Annotation Notes:**

1. For nested case-control studies and case-cohort studies, select "Yes" for both cohort study and case-control study
2. If the study is neither a cohort study nor a case-control study, select the appropriate option under other study design.
3. Please use the comment box sparingly. You may use it to:
4. Specify the study design if "Other" is selected, or
5. Note if the article appears to misclassify its study design.
6. Please use your own judgment rather than relying solely on the study's self-reported design.

### 3. Case Reports

Because the official MeSH definition is overly brief and general ("Clinical presentations that may be followed by evaluative studies that eventually lead to a diagnosis") (https://meshb.nlm.nih.gov/record/ui?ui=D002363), we chose a more detailed and explanatory definition, together with additional notes, taken from College of Human Medicine, Michigan State University [1]: A medical case report, also known as a case study, is a detailed description of a clinical encounter with a patient. The most important aspect of a case report, i.e. the reason you would go to the trouble of writing one, is that the case is sufficiently unique, rare or interesting such that other medical professionals will learn something from it.

**Notes to assist annotators in making a decision:**

1. Case reports are commonly of the following categories:

- Rare diseases
- Unusual presentation of disease
- Unexpected events
- Unusual combination of diseases or conditions
- Difficult or inconclusive diagnosis
- Treatment or management challenges
- Personal impact
- Observations that shed new light on a disease or condition
- Anatomical variations

It is important that you recognize what is unique or interesting about your case, and this must be described clearly in the case report.

2. Case reports generally take the format of:

- Background

- Case presentation
- Observations and investigation
- Diagnosis
- Treatment
- Outcome
- Discussion

**Additional notes:**

1. Case reports are generally incidental observations, not pre-planned studies. Case reports can report the results of interventions, but they should not be planned clinical trials.
2. Can be more than one patient, but generally a few patients, not a large series.
3. There can be overlap between "case report" and "case series" (see definition of case series [2]) so the two are not mutually exclusive.
4. Case reports can also be related not to patients, but to a group's experience with designing, rolling out, and evaluating software, hospital program roll-outs, and other such things. These are not CLINICAL case reports-- but they are also commonly called case reports. We marked non-clinical case reports as “UNSURE-nonclinical”.

### 4. Cross-Sectional Studies

Definition taken directly from the official MeSH definition (https://meshb.nlm.nih.gov/record/ui?ui=D003430):

Studies in which the presence or absence of disease or other health-related variables are determined in each member of the study population or in a representative sample at one particular time.

**Notes to assist annotators in making a decision:**

1. If the study involves one point in time, but does NOT measure each member of the study population nor a representative sample, this should be notated as "Cross-Sectional Design but not Cross-Sectional Study". (In practice, we found that neither NLM nor the automated taggers made the distinction between cross-sectional design and cross-sectional study, but marked both as Cross-Sectional Design. Often a survey was involved, which comprised a convenience sample and not a truly representative sample of the population.)
2. Cross-sectional studies contrast with LONGITUDINAL STUDIES which are followed over a period of time.
3. Additional notes taken from Capili [3]:

   Some related terms that point to cross-sectional studies include:

   - Disease Frequency Survey
   - Prevalence Study
4. Cross-sectional designs help determine the *prevalence* of a disease, phenomena, or opinion in a population, as represented by a study sample. *Prevalence* is the proportion of people in a population (sample) who have an attribute or condition at a specific time point regardless of when the attribute or condition first developed. Additionally, each study participant's evaluation is completed at one time-point with no follow-ups, providing a 'snapshot' of the sample.
5. A time point can actually be a period of time (e.g. a twelve month period) and not a single day.
6. Cross-sectional designs can be implemented as an interview or survey and may also collect physiological data and biological samples.
7. Cross-sectional studies can be descriptive or analytic.
8. Descriptive cross-sectional studies characterize the *prevalence* of health outcomes or phenomena under investigation. *Prevalence* is measured either at a one-time point (*point prevalence*), over a specified period (*period prevalence*) , or as a cross-sectional serial survey . The descriptive design starts by identifying the population of interest, collects the data, and classifies the participant, either as having the outcome or phenomena of interest or not.

9. Analytic cross-sectional studies can provide the groundwork to infer preliminary evidence for a causal relationship. This design allows investigators to identify a population or sample and collect *prevalence* data to evaluate outcome differences between exposed and unexposed participants on a disease, phenomena, or opinion. This design compares the proportion of participants exposed to the disease or phenomena of interest with the proportion of participants non-exposed with the disease or phenomena of interest. However, determining which variable is the dependent and independent variable or cause and effect is difficult to determine.
10. The *prevalence odds ratio (POR)* (calculated as [*ad/bc*]) and *prevalence ratio (PR)* (calculated as [a/(a + b)]/ [c/(c + d)]) are commonly used to report estimates of association between independent and dependent variables in cross-sectional studies.

### References


1 Writing a case report. https://research.chm.msu.edu/students-and-residents/writing-a-case-report (accessed 24 May 2026)

2 Shahidehpour A, Holt AW, Troy AM, *et al.* Creating an indexing scheme for case series articles. *medRxiv*. 2025;medRxiv:2025.12.19.25342712. doi: 10.64898/2025.12.19.25342712

3 Capili B. Cross-Sectional Studies. *Am J Nurs*. 2021;121:59–62. doi: 10.1097/01.naj.0000794280.73744.fe
